# Epithelial-endothelial barrier dysfunction and oxidative stress in ARDS subphenotypes: comparative efficacy of extracellular vesicles derived from bone marrow (MSC-EVs) and adipose tissue (ASC-EVs) mesenchymal stem cells

**DOI:** 10.64898/2026.09.22.26363677

**Authors:** Burello Alessandro, Tiberi Giorgia, Villegas Paula Ariadna Diez, Scagliotti Alessandro, Gai Chiara, Kopecka Joanna, Scrivanti Simone, Cappello Paola, Riganti Chiara, Brazzi Luca, Camussi Giovanni, Del Sorbo Lorenzo, Reddy Kiran, O’Kane Cecilia Majella, Cantaluppi Vincenzo, McAuley Danny Francis, Fanelli Vito

**Affiliations:** Department of Surgical Sciences, University of Turin, Italy; Department of Molecular Biotechnology and Health Sciences, Molecular Biotechnology Center “Guido Tarone”, University of Turin, Turin, Italy; Department of Medical Sciences, University of Turin, 10126 Turin, Italy; Department of Oncology, University of Turin, 10126 Turin, Italy; Interdepartmental Division of Critical Care Medicine, Department of Medicine, University of Toronto, Toronto, Ontario, Canada; Division of Respirology and Critical Care, Department of Medicine, University Health Network, Toronto, Ontario, Canada; Wellcome-Wolfson Institute for Experimental Medicine, Queen’s University of Belfast, Belfast, United Kingdom; Department of Critical Care, Belfast Health and Social Care Trust, Belfast, United Kingdom; Nephrology and Kidney Transplantation Unit, Department of Translational Medicine (DIMET), University of Piemonte Orientale (UPO), “Maggiore della Carità” University Hospital, Novara, Italy; Department of Anesthesia and Critical Care Medicine - Città della Salute e della Scienza University Hospital, Turin, Italy

**Keywords:** hyperinflammatory, hypoinflammatory subphenotypes, ARDS, ECMO, mesenchymal stem cell, extracellular vesicles

## Abstract

**Background:** Reactive oxygen species (ROS) overproduction, inflammatory cytokines, and plasma mediators disrupt alveolar tight junction integrity and perpetuate oxidative injury in acute respiratory distress syndrome (ARDS). Biological heterogeneity of ARDS, reflected by hyper- and hypoinflammatory subphenotypes with distinct plasma mediator profiles, may contribute to heterogeneity of treatment effect. Stem cell-derived EVs have been shown to exert regenerative and immunomodulatory effects by horizontal transfer of proteins, lipids and genetic material to injured target cells. This study investigated whether extracellular vesicles derived from bone marrow (MSC-EVs) and adipose tissue (ASC-EVs) mesenchymal stem cells restore barrier integrity and redox homeostasis, and whether efficacy differs across subphenotypes.

**Methods:** Bronchial epithelial cells (BEAS-2B), microvascular endothelial cells (HMEC-1), and small airway epithelial cells (SAEC) were challenged with plasma from patients with hyperinflammatory or hypoinflammatory ARDS subphenotypes, including those treated with venovenous ECMO. MSC-EVs or ASC-EVs were added concomitantly with plasma exposure or six hours later, at a dose of 5,000 particles/cell. After 24 h, the expression of actin and tight junction proteins, zonula occludens (ZO)-1, occludin, and vascular endothelial (VE)-cadherin, was evaluated by immunofluorescence. Oxidative stress parameters, antioxidant defenses, and antioxidant enzyme activities, were measured.

**Results:** Plasma from patients with the hyperinflammatory ARDS subphenotype induced marked disruption of ZO-1, occludin, and VE-cadherin expression, together with actin cytoskeletal rearrangement, across BEAS-2B, HMEC-1, and SAEC cells. Barrier disruption was accompanied by increased oxidative stress markers and altered antioxidant enzyme activity. Both MSC-EVs and ASC-EVs restored tight junction protein expression and cytoskeletal organization when administered either concurrently with ARDS plasma or 6 hours after exposure, and both effectively modulated redox parameters. Delayed administration 6 hours after injury produced greater effects for several oxidative stress endpoints, particularly in cells challenged with hypoinflammatory plasma.

**Conclusion:** ARDS-associated plasma mediators induce subphenotype-dependent impairment of epithelial-endothelial barrier integrity through oxidative mechanisms. Both MSC-EVs and ASC-EVs mitigate this injury, with efficacy that varies according to ARDS subphenotype and timing of administration.

## INTRODUCTION

The onset and progression of ARDS involves excessive ROS production by activated neutrophils and macrophages [1,2], which is further amplified by circulating plasma components,including cytokines such as IL-6 and IL-8, and hydroxyl radicals generated from cell-free hemoglobin [3]. Collectively, this oxidative stress disrupts alveolar barrier integrity through disassembly of tight junction proteins ZO-1, occludin, and VE-cadherin and actin cytoskeleton rearrangement, compromising the alveolar - capillary barrier [4,5]. Many of the inflammatory mediators that drive oxidative stress are also key biomarkers used to stratify patients with ARDS into two distinct biological subphenotypes: hypoinflammatory and hyperinflammatory. The hyperinflammatory subphenotype is characterized by elevated plasma levels of IL-6, IL-8, TNF-α, sTNFR-1, and Ang-2, several of which directly promote ROS generation and is associated with significantly higher mortality [6,7]. These subphenotypes respond differently to clinical interventions, underscoring the relevance of subphenotype-guided treatment strategies in ARDS [6–8]. Mesenchymal stem cells (MSCs) mitigate oxidative stress through several mechanisms, including the secretion of antioxidant enzymes, such as superoxide dismutase 3 (SOD3) and catalase, the transfer of functional mitochondria to damaged cells [9–11] and the release of extracellular vesicles carrying antioxidant enzymes and microRNAs [12]. These protective effects extend to the restoration of glutathione levels and modulation of key antioxidant enzymes [13]. However, the extent to which circulatory milieu from hyper- and hypoinflammatory ARDS patients induces oxidative stress and barrier disruption in pulmonary cells remains poorly understood, particularly with respect to the temporal dynamics of redox enzyme activation and tight junction protein expression [14,15]. Furthermore, althought MSC-EVs have shown therapeutic potential in experimental lung injury, comparative data on ASC-EV efficacy in ARDS are limited. Whether these two EV sources differentially restore barrier integrity and redox homeostasis across hyper- and hypo-inflammatory subphenotypes remains unknown [16,17]. This study examined the differential impact of hyper- and hypo-inflammatory ARDS plasma on epithelial- endothelial barrier integrity, and comparatively evaluated the timing-dependent therapeutic efficacy of MSC- and ASC-derived EVs in restoring barrier function and redox homeostasis.

We hypothesized that plasma from hyperinflammatory ARDS patients impairs barrier integrity by downregulating tight junction proteins, disrupting cytoskeletal organization and increasing oxidative stress. and that the distinct inflammatory microenvironments established by each subphenotype would differentially modulate the efficacy of MSC- and ASC-derived EVs.

## METHODS

A cohort of twenty patients with ARDS who had been determined to have a hyper- or hypoinflammatory subphenotype was included from an observational cohort study in the UK and Ireland [18]. Data and plasma samples were also collected from a second cohort of ten consecutive adult patients who were supported with VV ECMO for severe ARDS due to confirmed (real-time RT-PCR on nasopharyngeal swabs, or lower respiratory tract aspirates) COVID-19 (see inclusion/exclusion criteria and baseline characteristics data in online supplement) at the ECMO referral centre of the teaching hospital Città della Salute e della Scienza Turin. Blood was collected within 72 hour from ECMO placement, except for a patient in whom it was collected after 10 days. Internal review board of the hospital approved the study (protocol number 0028437). Plasma samples from five healthy volunteers served as controls. Whole blood was collected in lithium-heparin coated tubes (BD Vacutainer, Plymouth, UK). Plasma was separated from other components by centrifugation, then stored at −80°C, as previously described [18]. .

### Phenotyping of ARDS patients

ARDS patients were previously classified as hyperinflammatory or hypoinflammatory by a parsimonious model incorporating IL-6, sTNFR1, and bicarbonate [19]. This was done both prospectively using a near patient assay at the time of recruitment to the published PHIND study [18] and retrospectively in batch using gold standard Quantikine ELISA. Plasma samples were selected for extreme examples (probability >= 0.95 for hyperinflammatory patients and <= 0.05 for hypoinflammatory patients).

### Cell Cultures

BEAS-2B human bronchial epithelial cells and HMEC-1 human dermal microvascular endothelial cells were obtained from ATCC (Manassas, VA, USA), while SAEC human small airway epithelial cells were from Lonza (Walkersville, MD, USA). BEAS-2B cells were cultured in RPMI 1640 medium (PAN-Biotech) with 1% penicillin/streptomycin, 2 mM L-glutamine, and 10% heat-inactivated FBS (Gibco). HMEC-1 cells were grown in MCDB131 medium (Gibco) with 10 ng/ml EGF (Peprotech), 1 µg/ml hydrocortisone (Sigma Aldrich), 10 mM L-glutamine, 0.2% Mycozap Plus-CL (Lonza), and 10% heat-inactivated FBS. SAEC were cultured in Clonetics™ SAGM™ BulletKit™ (Lonza), including SABM™ Medium with growth supplements such as BPE, hydrocortisone, hEGF, epinephrine, transferrin, insulin, retinoic acid, triiodothyronine, gentamicin/amphotericin-B, and BSA-FAF.

### Extracellular vesicles (EVs) Isolation and Characterization

MSC-EVs and ASC-EVs were isolated from human bone marrow and adipose-derived Mesenchymal Stromal Cells (MSCs and ASCs) purchased from Lonza and cultured as described [20]. EVs were obtained by washing MSCs and ASCs at 70% confluence with PBS, then culturing in serum-free DMEM overnight at 37°C with 5% CO₂. Supernatants were centrifuged at 4000 rpm for 10 min at 4°C, filtered (0.22 μm), and ultracentrifuged twice at 100,000 rcf for 2 hours. Pellets were resuspended in PBS with 1% DMSO and stored at −80°C. EV concentration and size were assessed via nanoparticle tracking analysis (NTA) using a NanoSight LM10 system (405 nm laser, NTA 3.1 software) (Figure 3S A-B). Flow cytometry characterized EVs using FITC- or PE-conjugated antibodies (CD73, CD105, CD44), with isotypic IgG controls. The MACSPlex Exosome Kit (Miltenyi Biotec) was used for multiplexed bead-based analysis of 39 exosomal markers, counterstained with APC-conjugated antibodies (CD9, CD63, CD81), and analyzed on a CytoFLEX flow cytometer (Figure 3S C). EV integrity and morphology were analyzed by transmission electron microscopy (TEM) after glutaraldehyde fixation and negative staining (Figure 3S D). All experiments included three biological replicates. See online data supplement for methods details.

### Immunofluorescence and Image Quantification

BEAS-2B, HMEC-1, and SAEC cells were seeded on coated 8-well chamber slides at densities of 150,000, 90,000, and 70,000 cells/well, respectively. Once reaching 90-100% confluence, cultures were exposed to plasma from healthy volunteers and thirty ARDS patients, including ten treated with VV-ECMO and twenty previously classified as hyperinflammatory or hypoinflammatory by both prospective near patient assay and retrospective Quantikine ELISA gold standard, and select for extreme examples (probability >= 0.95 for hyperinflammatory patients and <= 0.05 for hypoinflammatory patients (1:4 dilution for BEAS-2B and SAEC, 1:10 for HMEC-1) [18] . A Cytomix solution 50 ng/mL (a mixture of human IL-1β, TNF-α, and IFN-γ (PeproTech, USA)) was included as a positive control [21]. MSC-EVs or ASC-EVs (5,000 EVs/cell) were added simultaneously or after 6 hours. After 24 hours, cells were washed, fixed in 4% paraformaldehyde, permeabilized with 0.2% Triton X-100, and blocked with 1% BSA. Primary antibodies (ZO-1, occludin, or VE-cadherin) were applied for 1 hour, followed by secondary antibodies (Cy3™ anti-rabbit, fluorescein anti-mouse) or Phalloidin-Tetramethylrhodamine B isothiocyanate for actin staining (Figure 1S). Slides were washed, dried and mounted with SlowFade™ Diamond Antifade Mountant with DAPI or TO-PRO™-3 Iodide. Images from the first cohort of samples were captured using a Leica TCS SP5 confocal system (405 nm diode, argon ion, 561 nm DPSS lasers, 63×/1. 4 NA oil objective, 0.24×0.24 μm resolution for BEAS-2B) or an Axiovert 200M LSM5 Pascal confocal microscope (40×/1.3 NA oil objective, 0.44×0.44 μm resolution for HMEC-1 and SAEC). Images from the second cohort of samples were acquired using an Agilent BioTek Cytation 5 in fluorescence mode (LED cubes, 20×/0.45 NA air objective), with DAPI (Ex 377/Em 447 nm) and RFP (Ex 531/Em 593 nm) filters, 0.32 µm resolution. Fluorescence images were captured using DAPI (Ex 377 nm/Em 447 nm) and RFP (Ex 531 nm/Em 593 nm) filter. The pixel size was 0.32 µm (3.12 pixels/µm). Fluorescence signals (ZO-1, Occludin, VE-Cadherin, Phalloidin-TRITC) were quantified as the Mean Gray Value, averaged from 3-5 images per well. Experiments were performed in duplicate. Image analysis was conducted using ImageJ software (NIH, Bethesda, MA, USA). See online data supplement for methods details

**Figure 1.**
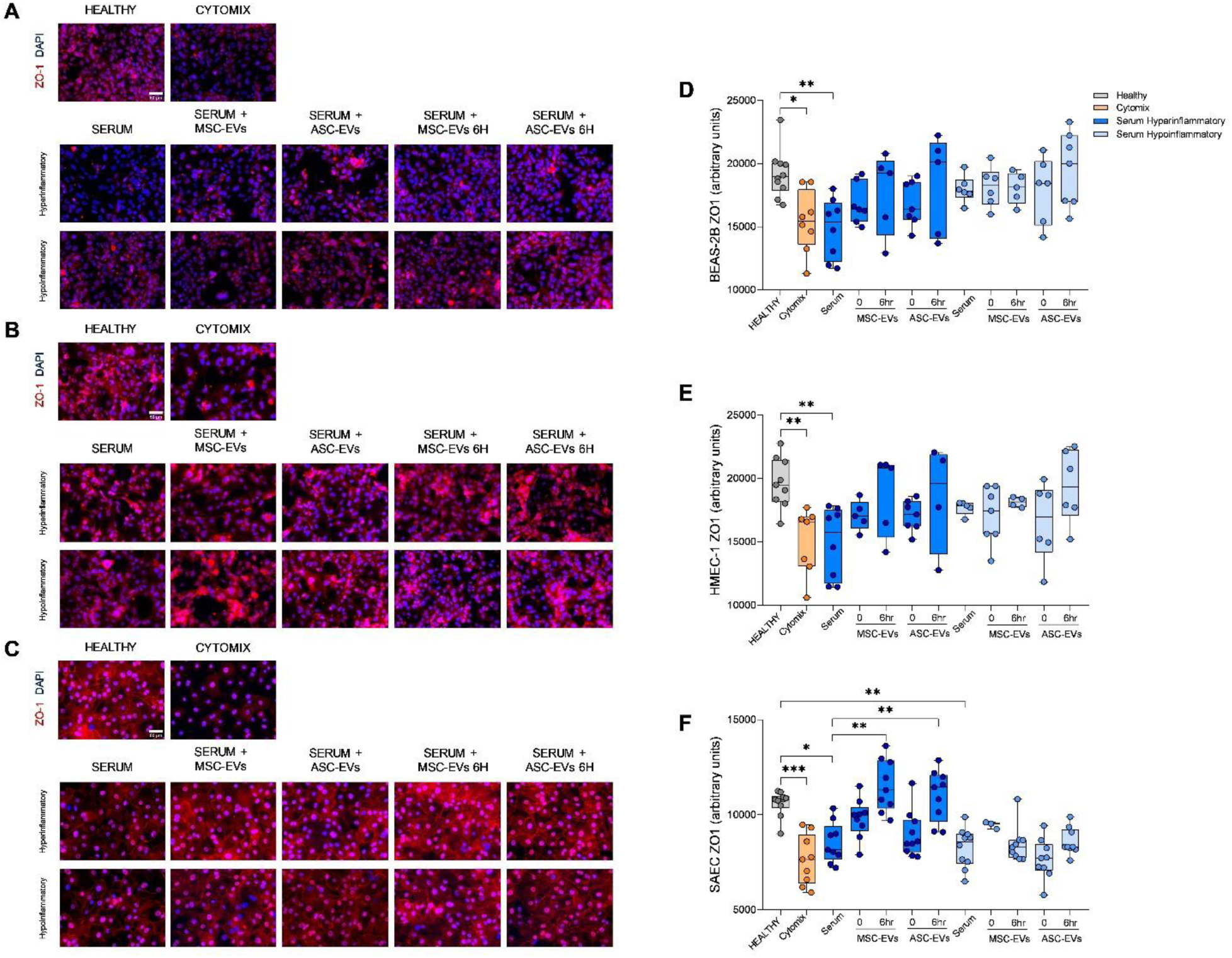
Representative images of immunofluorescence analysis (A) and quantitative analysis (D) of ZO-1 (red) expression in BEAS-2B cells, either challenged or unchallenged with MSC-EVs and ASC-EVs, concurrent with or 6 hours following exposure to plasma from ARDS patients with hyper- or hypoinflammatory subphenotypes, healthy volunteers or Cytomix. Immunofluorescence (B) and quantitative analysis (E) of ZO-1 (red) expression in HMEC-1 cells. Immunofluorescence (C) and quantitative analysis (F) of ZO-1 (red) expression in SAEC cells. The data are expressed as median and IQR. Individual points represent single measurements. Statistical significance was denoted as follows: *P<0.05, **P<0.01. Scale bar: 50 µm. Plasma tested: healthy, n = 5; hyperinflammatory, n = 10; hypoinflammatory subphenotype, n = 10.

### Oxidative stress

BEAS-2B, HMEC-1, and SAEC cells were seeded in 24-well plates at 120,000, 90,000, and 70,000 cells/well, respectively. At 90-100% confluence, cultures were exposed to plasma from healthy controls, ARDS patients (VV-ECMO (n =10), hyperinflammatory (n = 10) and hypoinflammatory (n = 10) subgroups; 1:20 dilution) or to Cytomix (50 ng/mL). MSC-EVs or ASC-EVs (5,000 particles/cell) were added simultaneously or after 6 hours (Figure 2S). Oxidative stress analyses for hyperinflammatory and hypoinflammatory ARDS subgroups were performed only in BEAS-2B and HMEC-1 cells. After 24 hours, cells were collected for oxidative stress assays. For ROS assays, 1×10⁵ cells were treated with 5 µM CM-H2DCFDA or MitoSOX for total and mitochondrial ROS detection. Malonyl dialdehyde (MDA) and protein carbonyl content were quantified using Abcam kits. Glutathione (GSH, GSSG) levels were measured after protein precipitation, with absorbance recorded at 415 nm. Glucose-6-phosphate dehydrogenase (G6PD) activity was assessed spectrophotometrically at 340 nm, differentiating total and 6-phosphogluconate dehydrogenase (6PGD) activity. Superoxide dismutase (SOD1, SOD2) activity was quantified via cytochrome c reduction at 550 nm. Glutathione reductase (GR), glutathione peroxidase (GPX), and thioredoxin reductase (TrxR) activities were measured using Abcam kits, with results expressed as enzymatic units/mg protein. See online data supplement for methods details.

**Figure 2.**
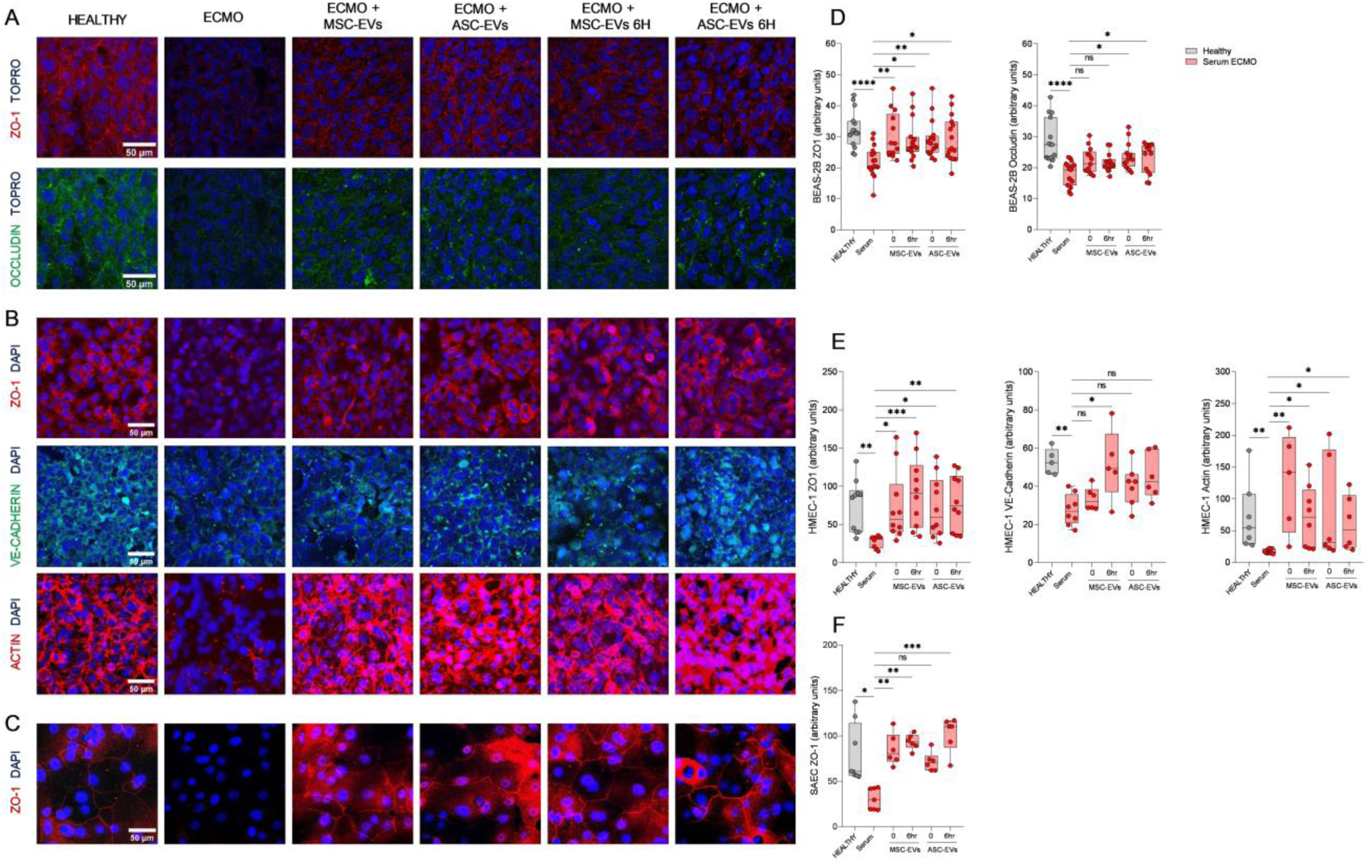
Representative images of immunofluorescence analysis (A) and quantitative analysis (D) of ZO-1 (red) and occludin (green) expression in BEAS-2B cells, either challenged or unchallenged with MSC-EVs and ASC-EVs, concurrent with or 6 hours following exposure to plasma from ARDS patients under vv ECMO or healthy volunteers. Immunofluorescence (B) and quantitative analysis (E) of ZO-1 (red), VE-cadherin (green), and actin (red) expression in HMEC-1 cells. Immunofluorescence (C) and quantitative analysis (F) of ZO-1 (red) expression in SAEC cells. The data are expressed as median and IQR. Individual points represent single measurements. Statistical significance was denoted as follows: *P<0.05, **P<0.01, ***P<0.001, ****P<0.0001. Scale bar: 50 µm. Plasma tested: healthy, n = 5; ECMO, n = 10.

**Figure 3.**
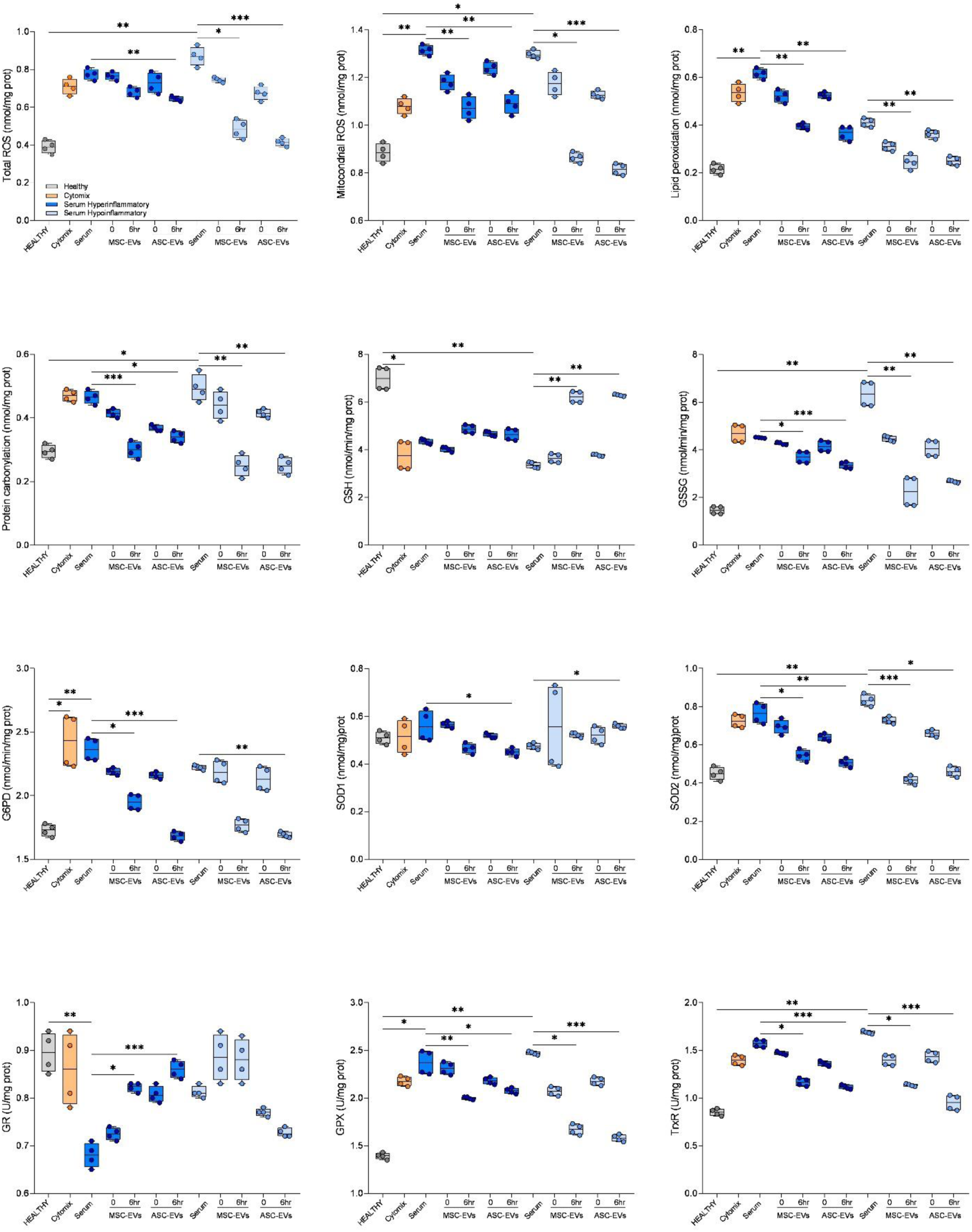
The concentrations of total reactive oxygen species (ROS), mitochondrial ROS (mtROS), lipid peroxidation, carbonylated proteins, reduced glutathione (GSH), oxidized glutathione (GSSG) and the activity of antioxidant enzymes, including glucose 6-phosphate dehydrogenase (G6PD), cytosolic superoxide dismutase 1 (SOD1), mitochondrial superoxide dismutase 2 (SOD2), glutathione reductase (GR), glutathione peroxidase (GPX), and thioredoxin reductase (TrxR) were measured in HMEC-1 cells following exposure to mesenchymal stem cell-derived extracellular vesicles (MSC-EVs) and adipose-derived stem cell extracellular vesicles (ASC-EVs), either concurrently or six hours post-exposure to plasma from ARDS patients with hyper or hypoinflammatory subphenotype, healthy volunteers or Cytomix. The data are expressed as median and IQR. Individual points represent single measurements. Statistical significance was denoted as follows: *P<0.05, **P<0.01. The plasma samples tested included those from healthy individuals (n = 5), hyperinflammatory ARDS patients (n = 10) and hypoinflammatory ARDS patients (n = 10).

### Statistical Analysis

Data were expressed as median and interquartile range (IQR) or proportions, as appropriate. The Kruskal-Wallis test followed by Dunn’s multiple comparison test was used for between-group comparisons. All statistical tests were two-tailed, and p-values less than 0.05 were considered statistically significant. Statistical analyses were performed using GraphPad Prism 8.0.1 (GraphPad, San Diego, CA, USA).

## RESULTS

### ARDS subphenotype classification

The baseline characteristics of patients are summarized in Table 1. Cells were challenged with plasma from ARDS patients previously classified as hyperinflammatory or hypoinflammatory by both prospective near patient assay and retrospective Quantikine ELISA gold standard, and selected for extreme examples (probability >= 0.95 for hyperinflammatory patients and <= 0.05 for hypoinflammatory patients) [18]

**Table 1.** Baseline characteristics of patients with ARDS.

| Variables | VV-ECMO<br>n = 10 | Hyperinflammatory<br>n = 10 | Hypoinflammatory<br>n = 10 |
| --- | --- | --- | --- |
| Age, years | 57 [49-59] | 59 [50-69] | 49 [45-59] |
| Gender Male, n (%) | 6 (60) | 2 (20) | 3 (30) |
| BMI, kg/m<sup>2</sup> | 29 [25-32] | 28 [23-31] | 30 [25-41] |
| Underlying comorbidities, n (%): |  |  |  |
| Obesity, | 6 (60) | 0 (0) | 2 (20) |
| Arterial hypertension, | 3 (30) | 2 (20) | 1 (10) |
| Hypothyroidism, | 2 (20) | 0 (0) | 0 (0) |
| SOFA, | 11 [10-13] | 19 [16, 20] | 9 [8, 9] |
| APACHE II | 21 [21-24] | 21 [18-26.8] | 14.5 [13-18.2] |
| Rescue therapies, n (%) |  |  |  |
| Lung recruitment maneuvers, n (%) | 2 (20) | 8 (80) | 5 (50) |
| Prone position, n (%) | 10 (100) | 2 (20) | 0 (0) |
| Inhaled nitric oxide, n (%) | 3 (30) | 2 (20) | 0 (0) |
| Ventilation settings at admission, |  |  |  |
| PaO<sub>2</sub>/FiO<sub>2</sub>, mmHg | 90 [80-100] | 121 [85-160] | 141 [106-149] |
| PaCO<sub>2</sub>, mmHg | 46 [40-48] | 37 [36-48] | 49 [44-63] |
| TV (ml/PBW) | 6.06 [5.77-6.19] | 8.4 [6.9-9.7] | 5.3 [4.4-7.1] |
| PEEP, cmH<sub>2</sub>O | 14 [12-16] | 8 [6-10] | 7 [5-9] |
| Plateau pressure, cmH<sub>2</sub>O | 29 [28-30] | 26 [21-31] | 28 [26-29] |
| Compliance, ml/cmH<sub>2</sub>O | 29 [26-30] | 26 [13-41] | 20 [19-21] |

Data are expressed as median [IQR] or n (%).
List of abbreviations: APACHE II = Acute Physiologic Assessment and Chronic Health Evaluation II; BMI = Body Mass Index; FiO<sub>2</sub> = Fraction of Inspired Oxygen; PaCO<sub>2</sub> = Partial pressure of arterial carbon dioxide; PaO<sub>2</sub> = Partial pressure of arterial oxygen; PEEP = Positive End-Expiratory Pressure; SOFA = Sequential Organ Failure Assessment score; TV = Tidal Volume; VV-ECMO = Veno-Venous Extracorporeal Membrane Oxygenation.

### Loss of tight junction protein expression and actin rearrangement induced by plasma from patients with ARDS

Three distinct cell lines, namely BEAS-2B, HMEC, and SAEC, were challenged with plasma from patients with hyperinflammatory and hypoinflammatory ARDS subphenotypes to assess the behavior of epithelial and endothelial barriers (Figure 1). Compared with healthy control plasma, ARDS plasma reduced ZO-1 expression across all cell types. The reduction was more pronounced in BEAS-2B and HMEC-1cells exposed to hyperinflammatory plasma than in those exposed to hypoinflammatory plasma (Figure 1A-E). In SAEC cells, both subphenotypes induced a similar reduction in ZO-1 (Figure 1C, F). Exposure to plasma from patients with severe ARDS treated with VV-ECMO caused a marked reduction in tight junction proteins expression in all three cell lines . Specifically, BEAS-2B cells showed decreased levels of ZO-1 and Occludin in comparison to cells exposed to plasma from healthy individuals (Figure 2A, D). HMEC-1 cells showed reduced ZO-1 and VE-cadherin expression, together with cytoskeletal reorganization indicated by decreased actin staining (Fig. 2B, E). SAEC cells showed a reduction in ZO-1 expression alone (Figure 2C, F).

### MSC-EVs and ASC-EVs restored the expression of tight junction proteins

In SAEC cells, MSC-EVs or ASC-EVs administered 6 hours after exposure to hyperinflammatory plasma restored ZO-1 expression to levels comparable with those observed in healthy controls (Figure 1C, F). In cells exposed to plasma from patients with severe ARDS treated with VV-ECMO, MSC-EVs or ASC-EVs administered either concomitantly or 6 hours after plasma exposure restored ZO- 1, occludin, VE-cadherin and actin expression to level comparable with those observed after exposure to healthy comtrol plasma (Figure 2A-F). No significant differences were detected between concomitant EV administration and administration 6 hours after exposure to the injurious stimulus.

### Oxidative stress induced by plasma from patients with ARDS is attenuated by the administration of MSC-EVs or ASC-EVs

Oxidative stress measurements showed increased levels of total reactive oxygen species (ROS), mitochondrial ROS (mROS), lipid peroxidation, and protein carbonylation in HMEC-1 (Figure 3) and BEAS-2B (Figure 6S) cells exposed to plasma from patients with hyper- and hypoinflammatory ARDS subphenotypes. Compared with healthy controls plasma, both subphenotypes increased total ROS, mROS, lipid peroxidation, protein carbonylation, and oxided glutathione (GSSG), while reducing glutathione (GSH) levels and altering the activity of G6PD, SOD2, GPX, and TrxR to an extent similar to Cytomix (Figure 3). Glutathione reductase (GR) activity was modulated only by plasma from patients with the hyperinflammatory subphenotype. EV treatment attenuated total ROS, mROS, lipid peroxidation, protein carbonylation and GSSG, particularly when administered 6 hours after plasma exposure. This effect was more evident in cells treated with hypoinflammatory plasma (Figure 3). EVs also restored G6PD, SOD2, GPX, and TrxR activity when added 6 hours after challenge with plasma from either subphenotype (Figure 3). Similar findings were observed in BEAS-2B cells (Figure 6S). A similar protective effect was observed when all three cell lines were exposed to plasma from patients with ARDS treated with VV-ECMO, compared with plasma from healthy volunteers (Figure 4, 4S, 5S). MSC-EVs and ASC-EVs, particularly when adminiostered six hours after exposure to ARDS plasma, reduced oxidative stress to levels comparable with those observed in cells exposed to healthy control plasma (Figure 4, 4S, 5S). Exposure to plasma from VV-ECMO patients also reduced GSH and increased GSSG content, and altered the activities of SOD1, SOD2, GPX, and TrxR. GR activity increased in HMEC-1 and BEAS-2B cells (Figure 4, 4S) but remained unchanged in SAEC cells (Figure 5S). MSC-EVs and ASC-EVs, particularly when administered six hours after plasma exposure, restored antioxidant defense activity to levels similar to those observed in cells exposed to healthy control plasma (Figure 4, 4S, 5S).

**Figure 4.**
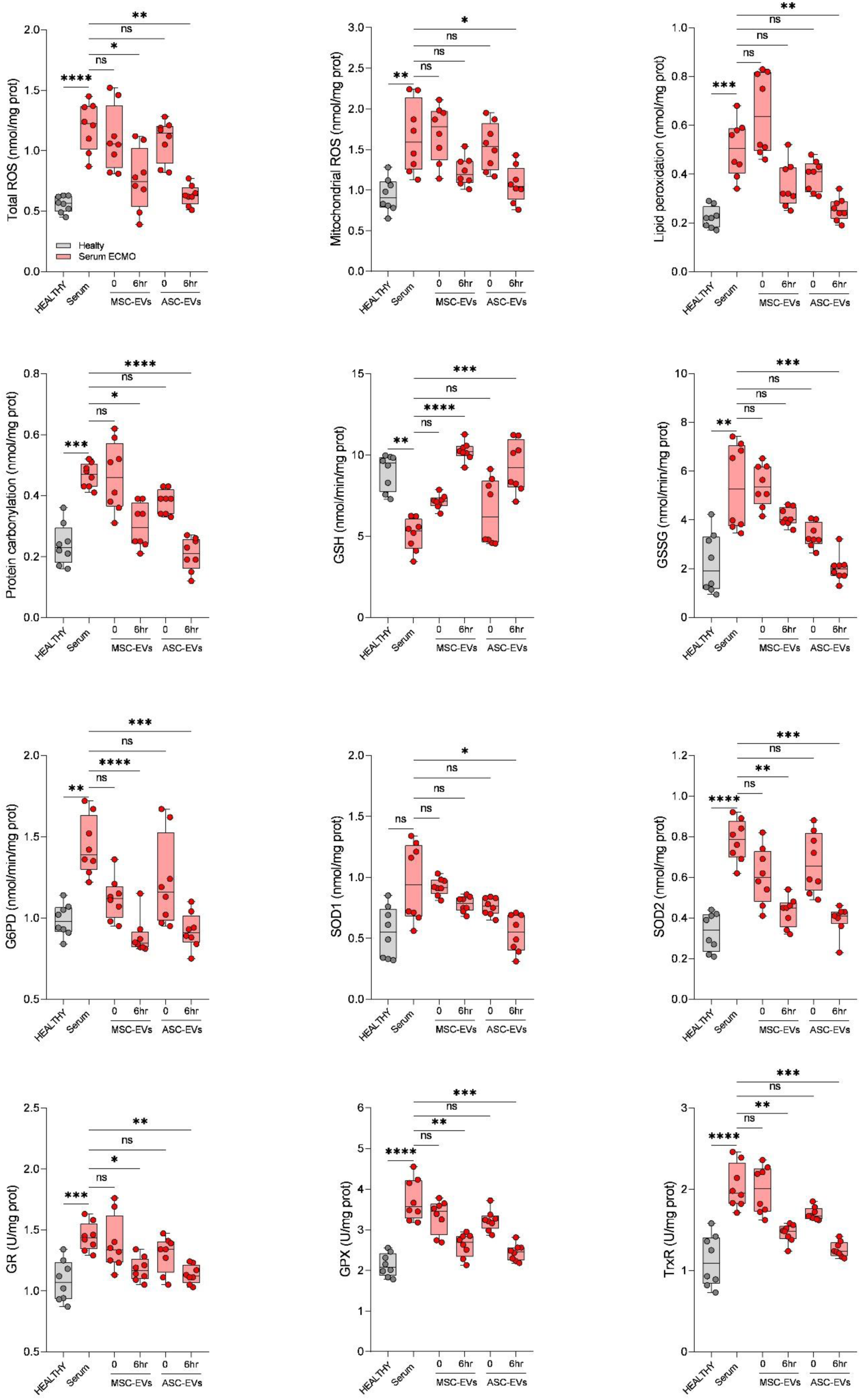
The concentrations of total reactive oxygen species (ROS), mitochondrial ROS (mtROS), lipid peroxidation, carbonylated proteins, reduced glutathione (GSH), oxidized glutathione (GSSG) and the activity of antioxidant enzymes, including glucose 6-phosphate dehydrogenase (G6PD), cytosolic superoxide dismutase 1 (SOD1), mitochondrial superoxide dismutase 2 (SOD2), glutathione reductase (GR), glutathione peroxidase (GPX), and thioredoxin reductase (TrxR) measured in HMEC-1 cells following exposure to mesenchymal stem cell-derived extracellular vesicles (MSC-EVs) and adipose-derived stem cell extracellular vesicles (ASC-EVs), either concurrently or six hours post-exposure to plasma from patients with ARDS under vv ECMO or healthy volunteers. The data are expressed as median and IQR. Individual points represent single measurements. Statistical significance was denoted as follows: *P<0.05, **P<0.01, ***P<0.001, ****P<0.0001. The plasma samples tested included those from healthy individuals (n = 5) and ARDS patients under vv ECMO (n = 10).

## DISCUSSION

The principal finding of this study is that both MSC-EVs and ASC-EVs restored barrier integrity and redox homeostasis in epithelial and endothelial cell exposed to ARDS plasma. Therapeutic efficacy was maintained, and in several oxidative stress endpoints enhanced, when EVs were administered 6 hours afterv injury, particularly in cells challenged with plasma from patients with hypoinflammatory subphenotype.

ARDS plasma-induced disruption of tight junction integrity highlights the complex interplay between inflammatory mediators, oxidative stress, and barrier function. Plasma from both hyper- and hypoinflammatory ARDS subphenotypes induced oxidative modifications of cellular lipids and proteins. The increase in lipid peroxidation and GPX activity are consistent with reports that oxidized phospholipids accumulate in ARDS, impair surfactant function, and promote cell death by ferroptosis [22,23]. Protein carbonylation, a marker of oxidative stress, has been shown to inhibit histone deacetylases (HDACs) and sirtuins, leading to aberrant gene expression, particularly of inflammatory mediators [24]. In our model, increased mROS production is consistent with evidence that mitochondrial dysfunction is an early event in mitochondrial ROS-dependent activation of the NLR family pyrin domain containing 3 protein (NLRP3) inflammasome during sepsis [25]. We also observed increased SOD1and SOD2 activity in epithelial and endothelial cells, suggesting an attempted compensatory antioxidant response to preserve barrier integrity [26]. However, despite increased activity of antioxidant enzymes such as SOD and GPX, these compensator responses were insufficient to restore redox homeostasis. The therapeutic efficacy of MSC-EVs and ASC-EVs extends previous findings on EV-based treatment in experimental pneumonia, in which EVs increased intracellular ATP levels in injured alveolar epithelial type 2 cells [27]. The sustained efficacy observed 6 hours after injury builds on prior work addressing the timing of mesenchymal stem cell interventions [28]. In previous studies, intratracheal EV administration reduced lung permeability and inflammation 48 hours after Escherichia coli endotoxin-induced lung injury, partly through the upregulation of keratinocyte growth factor (KGF) mRNA in injured alveolar tissue [28]. Several mechanisms may explain the beneficial effects of EVs. MSC-derived EVs can transfer functional mitochondria to injured type II alveolar epithelial cells [29] and alveolar macrophages, thereby improving alveolar–capillary barrier function [9,11,30]. In addituion, EVs may deliver reparative mediators such as keratinocyte growth factor (KGF), thereby promoting epithelial repair [28]. The more pronounced therapeutic response observed under hypoinflammatory conditions may reflect a differential receptivity of target cells to EV-mediated reparative mechanisms. Under the severe oxidative and inflammatory burden characteristic of the hyperinflammatory subphenotype, cellular capacity to integrate mitochondrial transfer, miRNA signaling, and growth factor delivery may be compromised, thereby limiting the therapeutic impact of EVs. Although our results demonstrate that EVs restore barrier function and redox balance, the precise molecular mechanisms underlying these effects require further investigation. Previous studies have reported that MSCs exhibit low basal ROS levels and high concentrations of glutathione, the major cellular antioxidant, at baseline. [31,32]. Conversely, other studies indicate that MSCs may have limited antioxidant capacity and greater vulnerability to oxidative stress than more differentiated cell types [31,33]. To our knowledge, this is the first study to compare MSC-EVs and ASC-EVs in ARDS plasma-induced barrier dysfunction. Unlike previous work focused primarily on MSC-EVs [17,28], our findings show comparable efficacy of ASC-EVs, thereby broadening the potential therapeutic relevance of EV-based strategies. The persistence of EV efficacy beyond 6 hours after injury also suggests a clinically relevant therapeutic window.

Several limitations should be considered when interpreting these findings. Firstly, the mechanism of EV action was not directly investigated; therefore, our conclusions are limited to their observed effects on epithelial and endothelial barrier integrity and oxidative stress. Second, although we demonstrated acute responses to ARDS plasma, sampling reflected a single time point in a dynamic disease process and may not capture the temporal evolution of inflammatory mediators, as shown in studies of bronchoalveolar lavage cytokine profiles [34]. Third, although EV characterization was comprehensive, it may not have captured all bioactive cargo components, and the long-term effects of oxidative stress on EV efficacy remains unknown [27]. Fourth, although the in vitro system used here demonstrates key therapeutic potential, clinical transition will require validation in more complex models that reproduce the heterogeneity of hyper- and hypoinflammatory ARDS. Fifth, the optimal timing and dosing of EV administration require validation in experimental settings that include the complex inflammatory milieu, immune-cell interactions, and mechanical forces present in ARDS.

In conclusion, our study demonstrates the therapeutic potential of MSC- and ASC-derived EVs in restoring barrier integrity and redox homeostasis. The enhanced efficacy observed under hypoinflammatory conditions suggests that the inflammatory microenvironment critically determines the extent of EV-mediated repair and identifies a potentially relevant therapeutic window at 6 hours after injury. Future study should characterize the EV cargo responsible for these subphenotype-dependent effects and evaluate EV-based strategies in more complex experimental models that recapitulate hyper- and hypoinflammatory ARDS.

## List of abbreviations

ECMO: extracorporeal membrane oxygenation
ARDS: acute respiratory distress syndrome
ICU: intensive care unit
RT-PCR: real time - polymerase chain reaction
PaO2: partial pressure of arterial oxygen
PaCO2: partial pressure of carbon dioxide
FiO2: fraction of inspired oxygen
BMI: body mass index
PBW: predicted body weight
SAPS II: Simplified Acute Physiology Score II
SOFA: sequential organ failure assessment
iNO: inhaled nitric oxide
LRM: lung recruitment manoeuvres
PEEP: positive end expiratory pressure
SARS-CoV-2: severe acute respiratory syndrome coronavirus 2

## Declarations

Ethics approval and consent to participate: Institutional review board approved the study protocol and waived the requirement for informed consent. Protocol number approval 0028437 for Turin ECMO center. For the hypo and hyper inflammatory cohorts, the protocol was approved by research ethics committees in England, Wales, Northern Ireland (19/LO/0672), Scotland (19/SS/0073), and Ireland (Ca. 2344). The study protocol was prospectively published online and was registered on ClinicalTrials.gov, NCT04009330. The study was conducted in accordance with Good Clinical Practice guidelines, local regulations, and the ethical principles described in the Declaration of Helsinki. Written informed consent was obtained from patients’ surrogates before recruitment. As this was deemed a low-risk study, when a surrogate decision-maker was not available, a deferred consent model was utilised in England, Wales, and Northern Ireland (but not in Scotland or Ireland). If deferred consent was not obtained, the patient was withdrawn from the study, samples were destroyed, and all data were removed. Written consent to continue participation in the study was sought for all patients who regained capacity following intensive care discharge [18].

All human cells used in the in vitro experiments were obtained from commercial suppliers.

BEAS-2B human bronchial epithelial cells (ATCC, CRL-9609) and HMEC-1 human dermal microvascular endothelial cells (ATCC, CRL-3243) are established, immortalised cell lines obtained from a public repository and are not primary human cells. BEAS-2B cells were originally derived from normal human bronchial epithelium obtained at autopsy from non-cancerous individuals and immortalised with an adenovirus 12-SV40 hybrid virus by the source laboratory [35]. HMEC-1 cells were originally derived from the dermal microvascular endothelium of neonatal human foreskin and immortalised by transfection with the SV40 large T antigen at the Centers for Disease Control and Prevention, and were deposited with ATCC in 1990 [36]. Both cell lines were used under the repository’s standard terms and conditions for laboratory research use only, and no further ethical approval was required for their use in this study.

SAEC human small airway epithelial cells (Lonza, CC-2547), human bone marrow-derived Mesenchymal Stromal Cells (Lonza, PT-2501) and human adipose-derived Mesenchymal Stromal Cells (Lonza, PT-5006) are primary human cells. According to the supplier, bone marrow-derived Mesenchymal Stromal Cells are isolated from bilateral punctures of the posterior iliac crests of normal adult volunteers, and adipose-derived Mesenchymal Stromal Cells are isolated from lipoaspirates collected during elective surgical liposuction procedures. The supplier has confirmed that all human tissue used to manufacture these products is ethically obtained from donors, or their legal representatives, who provided written informed consent under consent forms and protocols approved by an Institutional Review Board or an equivalent regulatory authority, and that the material is supplied with documented legal permission for research use. All cells were used in accordance with the supplier’s terms and conditions of sale for non-commercial research purposes, and no further ethical approval was required for the use of these commercially available cells.

## Consent for publication

Not applicable

## Availability of data and materials

The de-identified dataset supporting the findings of this study has been deposited in Zenodo and is publicly available at the following DOI: 10.5281/zenodo.22693238.

## Authors’ Contribution

Burello A: data interpretation and analysis, manuscript writing. Tiberi G: data interpretation and analysis, manuscript writing. Villegas PAD: data interpretation and analysis, manuscript writing. Scagliotti A: data interpretation and analysis, manuscript revision. Cappello P: study design, data interpretation and analysis, manuscript revision. Gai C: data interpretation and analysis, manuscript revision. Kopecka J: data interpretation and analysis, manuscript revision Riganti C: data interpretation and analysis, manuscript revision. Brazzi L: data interpretation and analysis, manuscript revision. Camussi G: data interpretation and analysis, manuscript revision. Del Sorbo L: data interpretation and analysis, manuscript revision. Reddy K: data interpretation and analysis, manuscript revision. O’Kane CM: data interpretation and analysis, manuscript revision. Cantaluppi V: data interpretation and analysis, manuscript revision McAuley DF: data interpretation and analysis, manuscript revision. Fanelli V (corresponding author): study design, data interpretation and analysis, manuscript writing and revision.

## Data Availability

All data produced are available online in Zenodo at the following DOI: 10.5281/zenodo.22693238.

## Aknowledgments

During the preparation of this manuscript, the authors used Paperpal exclusively for AI-assisted copy editing, that is, to improve the readability, grammar, spelling and style of text that had been entirely written by the authors. No generative AI tool was used to generate scientific content, to design the study, to produce, analyse or interpret the data, or to create or modify any figure or image. The authors reviewed and edited all AI-assisted text and take full responsibility for the content of the publication.

## Turin

The authors would like to thank Vincenzo Elia for technical support in the lab and all nurses and perfusionists involved in ECMO program.

