## Supplementary material for "Epithelial-endothelial barrier dysfunction and oxidative stress in ARDS subphenotypes: comparative efficacy of extracellular vesicles derived from bone marrow (MSC-EVs) and adipose tissue (ASC-EVs) mesenchymal stem cells": Supplemets

**ONLINE DATA SUPPLEMENT**

#### METHODS

##### Patients

A cohort of twenty patients with ARDS who had been determined to have a hyper- or hypo-inflammatory subphenotype was included from a multicentre, prospective, observational cohort study in the UK and Ireland (Reddy et al. 2026). ARDS was defined by the Berlin definition (Ards Definition Task Force et al. 2012). Exclusion criteria included: age younger than 18 years; more than 72 h after onset of ARDS or AHRF (which was amended during the course of the study from more than 48 h after onset of ARDS to facilitate patient recruitment and justified based on evidence supporting subphenotype stability16); receiving extracorporeal membrane oxygenation at the time of recruitment; treatment withdrawal imminent within 24 h; a do not attempt resuscitation order in place; anyone who declined consent; or people who were incarcerated (Reddy et al. 2026).

Data and plasma samples were also collected also from a cohort of ten consecutive adult patients who were supported with VV ECMO for severe ARDS due to confirmed (real-time RT-PCR on nasopharyngeal swabs, or lower respiratory tract aspirates) COVID-19 (see inclusion/exclusion criteria and baseline characteristics data in online supplement) at the ECMO referral centre of the teaching hospital Città della Salute e della Scienza di Torino. Internal review board of the hospital approved the study (protocol number 0028437). Patients were considered eligible for VV ECMO if under protective mechanical ventilation (tidal volume of 6 ml/kg to keep plateau pressure below 30 cmH2O), sedation, muscle paralysis had a ratio of partial pressure of arterial oxygen ($PaO_2$) to the fraction of inspired oxygen ($FiO_2$) less than 100 (Combes et al. 2018). Patients were considered not suitable for extracorporeal support if any of the following: injurious ventilation at plateau pressure > 30 $cmH_2O$ for more than a week, contraindication to systemic anticoagulation with heparin, chronic respiratory failure requiring oxygen therapy or non-invasive ventilation, cancer with a life expectancy of less than 5 years, moribund patient as judged by the treating physician and logistic situation in which the ECMO mobile service was not immediately available (Combes et al. 2018). Pump speed and $FiO_2$ on ECMO were adjusted to obtain blood-oxygen saturation of more than 90% with $FiO_2$ on ventilator <60%. To minimize the risk of ventilator induced lung injury sweep gas was adjusted to allow a tidal volume of 4 ml/kg and plateau pressure <25 cmH2O and driving pressure (plateau pressure-peep) <15 cmH2O (Combes et al. 2018; Patroniti et al. 2011). Anticoagulation with unfractionated heparin was adjusted to target a ratio of activated partial thromboplastin time between 1.51-2.34 (Combes et al. 2018; Patroniti et al. 2011). Baseline characteristics included dates of admission to the referral ICU, age, sex, weight and height used to calculate the body mass index (BMI) and the predicted body weight (PBW), presence of coexisting chronic disease, initial severity assessed by the SAPS II score and organ failures assessed by the SOFA score. Before ECMO initiation, we recorded the time and duration of hospitalization, the use of rescue therapies such as prone positioning, inhaled nitric oxide (iNO), lung recruitment manoeuvres (LRM), respiratory parameters (i.e. tidal volume, positive end expiratory pressure, plateau pressure, driving pressure, PaO2/FiO2 ratio), arterial blood gas.

**Cell Cultures**

BEAS-2B human bronchial epithelial cells and HMEC-1 human dermal microvascular endothelial cells were obtained from American Type Culture Collection (ATCC, Manassas, VA, USA). SAEC, human small airway epithelial cells, were obtained from the distal airspace (Clonetics™ Small Airway Epithelial Cell Systems, Lonza, Walkersville, MD, USA). BEAS-2B cells were cultured in Roswell Park Memorial Institute 1640 medium (RPMI1640, PAN-Biotech, Aidenbach, Germany) supplemented with 1% penicillin/streptomycin (Gibco, Thermo Fisher Scientific, Waltham, MA, USA), 2 mM L-glutamine (Gibco, Thermo Fisher Scientific, Waltham, MA, USA), and 10% v/v heat-inactivated Fetal Bovine Serum (FBS, Gibco, Thermo Fisher Scientific, Waltham, MA, USA). HMEC-1 were cultured in Molecular, Cellular, and Development Biology 131 medium (MCDB131, Gibco, Thermo Fisher Scientific, Waltham, MA, USA) supplemented with 10 ng/ml Epidermal Growth Factor (EGF, Peprotech), 1 µg/ml Hydrocortisone (Sigma Aldrich, Merck, Darmstadt, Germany), 10 mM L-Glutamine (Sigma Aldrich, Merck, Darmstadt, Germany), 0.2% of Mycozap Plus-CL (Lonza, Basel, Switzerland), 10% v/v of Heat inactivated Fetal Bovine Serum (FBS, Gibco, Thermo Fisher Scientific, Waltham, MA, USA). SAEC were cultured in Clonetics™ SAGM™ BulletKit™ (CC-3118) contains 500 ml of Small Airway Epithelial Cell Basal Medium (SABM™ Medium) and the following growth supplements: Bovine Pitituary Extract (BPE), 2 ml; Hydrocortisone, 0.5 ml; human Epidermal Growth Factor (hEGF), 0.5 ml; Epinephrine, 0.5 ml; Transferrin, 0.5 ml; Insulin, 0.5 ml; Retinoic Acid, 0.5 ml; Triiodothyronine, 0.5 ml; Gentamicin/Amphotericin-B, 0.5 ml; Bovine Serum Albumin – Fatty Acid Free (BSA-FAF), 5.0 ml (Lonza, Walkersville, MD, USA).

**EVs isolation and characterization**

MSC-EVs and ASC-EVs were isolated from human bone marrow Mesenchymal Stromal Cells (MSCs) and adipose Mesenchymal Stromal Cells (ASCs), respectively. Both cell lines were purchased from Lonza (Basel, Switzerland) and cultured as described previously (Pomatto et al. 2021). Briefly, EV were isolated as follows. MSCs and ASCs at 70% confluence were thoroughly washed with PBS (Lonza, Basel, Switzerland) to remove serum residues. They were then cultured overnight (16 hours) in DMEM (Lonza, Basel, Switzerland) supplemented withpenicillin/streptomycin and L-glutamine (Sigma-Aldrich, St. Louis, MO, USA), but without Fetal Bovine Serum (FBS) (Thermo Fisher Scientific, Waltham, MA, USA), under 5% $CO_2$ at 37°C. The cell culture supernatants were centrifuged at 4000 rpm for 10 min at 4°C and filtered through a 0.22 µm vacuum filter unit (Millipore,

Burlington, MA, USA) to remove cell debris and apoptotic bodies. They were subsequently ultracentrifuged twice at 100,000 rcf for 2 hours at 4°C using a Beckman Coulter Optima L-90K ultracentrifuge with a 45 Ti rotor and polycarbonate tubes (Beckman Coulter, Indianapolis, IN, USA). The resulting pellets were resuspended in PBS (Lonza, Basel, Switzerland) containing 1% DMSO (Sigma-Aldrich, St. Louis, MO, USA) and stored at -80°C for future experiments. EVs' concentration and size of EVs were analyzed by nanoparticle tracking analysis (NTA) using a NanoSight LM10 system (NanoSight, Salisbury, UK) as previously described (Pomatto et al. 2021). The NanoSight system was equipped with a 405 nm laser and NTA 3.1 analytic software. Briefly, EVs were diluted 1:200 in 1 mL of saline solution (Fresenius Kabi, Bad Homburg Vor der Höhe, Germany) that had been pre-filtered using 0.22 μm membranes (Millipore, Burlington, MA, USA). For each sample, three 30-second videos were recorded with the camera levels set to 15 for all acquisitions. The post-acquisition settings for NTA were optimized and maintained consistently across all samples. Each video was subsequently analyzed to determine the mean size and concentration of EVs. EVs' integrity and morphology of EVs were assessed using transmission electron microscopy. Briefly, fresh EV samples were stained with 2.5% glutaraldehyde containing 2% sucrose and negatively stained with Nano-W and NanoVan (Nanoprobes, Yaphank, NY, USA) as previously described (Pomatto et al. 2021). The preparations were analyzed using a Jeol JEM 1010 electron microscope (Jeol, Tokyo, Japan). EVs were characterized by cytofluorometric analysis using fluorescein isothiocyanate (FITC)-or phycoerythrin (PE)-conjugated antibodies targeting CD73, CD105, and CD44 (Miltenyi Biotec, Bergisch Gladbach, Germany). Conjugated mouse non-immune isotypic immunoglobulin G (IgG) (Miltenyi Biotec, Bergisch Gladbach, Germany) served as a control. Briefly, 10 μL of EVs were labeled with antibodies for 15 min at 4°C, diluted 1:3 immediately, and acquired. For analysis using the human bead-based MACSPlex Exosome

Kit (Miltenyi Biotec, Bergisch Gladbach, Germany), the manufacturer's protocol was followed. EV samples, containing approximately 1 × $10^9$ particles per preparation, were diluted in MACSPlex buffer (MPB) to a final volume of 120 μL in 1.5 mL microcentrifuge tubes. Each sample was mixed with 15 μL MACSPlex exosome capture beads (a cocktail of 39 exosomal marker epitopes) and counterstained with 15 μL APC-conjugated antibodies (anti-CD9, anti-CD63, and anti-CD81). Samples were incubated overnight at room temperature on an orbital shaker at 450 rpm in the dark. After incubation, the beads were washed with 1 mL of MPB by centrifugation at 3000 × g for 5 min, followed by a longer wash step with 1 mL of MPB, and then incubated on an orbital shaker for 15 min. The beads were centrifuged again at 3000 × g for 5 min and the supernatant was aspirated, leaving a residual volume of 150 μL per tube. During acquisition, the median fluorescence intensity (MFI) for all 39 exosomal markers was corrected for a medium background and gated according to the fluorescence intensity according to the manufacturer's instructions. Cytofluorometric analysis was performed using a CytoFLEX flow cytometer (Beckman Coulter, Indianapolis, IN, USA) equipped with CytExpert software, version 2.3.0.84. Both classical FACS and MACSPlex exosome kit analyses included three biological replicates.

**Immunofluorescence and Images Quantification**

BEAS-2B cells were seeded on Corning® BioCoat® Collagen I 8-well Culture Slide (Corning, Gleandale, Arizona, US) at a density of 150,000 cells/well, HMEC-1 cells were seeded in Lab-Tek Chamber Slidee 8 wells (Thermo Fisher Scientific, Waltham, MA, USA) at a density of 90,000 cells/well, and SAEC were seeded on Lab-Tek Chamber Slidee 8 wells (Thermo Fisher Scientific, Waltham, MA, USA) at a density of 70,000 cells/well. When the cells reached 90%-100% confluence, the culture was exposed to plasma from healthy volunteers or plasma from ARDS patients (VV-ECMO (n =10), hyperinflammatory (n =10) and hypoinflammatory (n = 10) subgroups) diluted in fresh growth medium 1:4 for BEAS-2B and SAEC or 1:10 for

HMEC-1. An additional control was included using Cytomix (50 ng/mL), a mixture of human IL-1β, TNF-α, and IFN-γ (PeproTech, USA), used as a surrogate for ARDS pulmonary edema fluid (Fang et al. 2010). MSC-EVs or ASC-EVs were added simultaneously to the plasma or after 6 h at a dose of 5,000 particles/cell. After 24 h, the cell monolayer was washed twice with Phosphate Buffered Saline (PBS) and fixed in 4% paraformaldehyde for 10 min. The cells were then washed thrice with PBS for 5 min and permeabilized with 0.2% Triton X-100 for 5 min. The slides were again washed thrice with PBS for 5 min and then blocked with 1% BSA for 30 min at room temperature. For primary antibody staining, the slides were incubated with Zonula occludens (ZO)-1 rabbit anti-human primary antibodies (Invitrogen, Thermo Fisher Scientific, Waltham, MA, USA) at a concentration of 7.5 µg/ml and occludin mouse anti-human primary antibodies (Invitrogen, Thermo Fisher Scientific, Waltham, MA, USA) at a concentration of 2.5 µg/ml or vascular endothelial (VE)-Cadherin Monoclonal Antibody BV9 (Invitrogen, Thermo Fisher Scientific, Waltham, MA, USA) at a concentration of 2.5 µg/ml for 1 h at room temperature. After washing with PBS three times, slides were incubated with secondary antibody Cy3™ goat anti-rabit IgG (H + L) (Invitrogen, Thermo Fisher Scientific, Waltham, MA, USA) and Fluorescein got anti-mouse IgG (H + L) (Invitrogen, Thermo Fisher Scientific, Waltham, MA, USA) at a concentration of 5 µg/ml, or, for Actin staining, with Phalloidin-Tetramethylrhodamine B isothiocynate (Sigma Aldrich, Merck, Darmstadt, Germany) at a concentration of 0.2 µg/ml for 1 hour at room temperature. The slides were then washed with PBS and dried at room temperature. Slides were then mounted using SlowFade™ Diamond Antifade Mountant with DAPI (Invitrogen, Thermo Fisher Scientific, Waltham, MA, USA) or TO-PRO™-3 Iodide (Invitrogen, Thermo Fisher Scientific, Waltham, MA, USA). Images from VV-ECMO samples were captured using a Leica TCS SP5 confocal system (Leica Microsystems) equipped with a 405 nm diode, an argon ion, a 561 nm DPSS lasers, with a 63×/1.4 NA oil immersion objective and acquired with a resolution of 0,24×0,24 µm for

BEAS-2B or with the confocal microscope Axiovert 200M equipped with LSM5 Pascal (Zeiss, Oberkochen, Germany) with a 40×/1.3 NA oil immersion objective and acquired with a resolution of 0,44×0,44 µm for HMEC-1 and SAEC. For experiments using hyperinflammatory and hypoinflammatory ARDS plasma, images were acquired using an Agilent BioTek Cytation 5 Cell Imaging Multi-Mode Reader CYT5FV in fluorescence imaging mode, equipped with LED cubes. A 20×/0.45 NA PL FL phase contrast objective (air, long working distance 6.4–7.6 mm) was used. Fluorescence images were captured using DAPI (Ex 377 nm/Em 447 nm) and RFP (Ex 531 nm/Em 593 nm) filter. The pixel size was 0.32 µm (3.12 pixels/µm). The fluorescence signals of ZO-1, Occludin, VE-Cadherin and Phalloidin-TRITC (Actin) staining were quantified as the Mean Gray Value, which is the sum of the gray values of all the pixels in the image divided by the number of pixels. For each well, the Mean Gray Value of 3-5 to images and the mean values were used for statistical analysis. Images were processed and analyzed using the ImageJ software (Rasband, W.S., USA). National Institutes of Health, Bethesda, MA), (Figure 1S).

**Oxidative stress assays**

BEAS-2B, HMEC-1, and SAEC cells were seeded in 24-well plates (Euroclone, Milan, Italy) at a density of 120, 000, 90, 000, and 70, 000 cells/well, respectively.

When the cells reached 90-100% confluence, the culture was exposed to plasma from healthy volunteers, plasma from ARDS patients (VV-ECMO (n = 10), hyperinflammatory (n = 10) and hypoinflammatory (n = 10) subgroups) diluted in fresh growth medium (1:20) or Cytomix (50 ng/mL). MSC-EVs or ASC-EVs were added simultaneously to plasma or after 6 h at a dose of 5,000 particles/cell. After 24 h, the supernatant was removed and cells were detached with TRY-EDTA (Gibco, Thermo Fisher Scientific, Waltham, MA, USA) and centrifuged as per the manufacturer's instructions to obtain dried pellets of approximately 1 × 106 cells per experimental condition.

For total and mitochondrial (mt)ROS assays, 1×105 cells were washed with PBS and detached by gentle scraping. A 50 µL aliquot was sonicated and used to measure cellular proteins. The remaining cells were treated for 30 min at 37 °C with 5µM µM ROS-sensitive fluorescent probes 5-(and-6)-chloromethyl-2’,7’-dichlorodihydro-fluorescein diacetate (CM-H2DCFDA) (ThermoFisher, Waltham, MA) or with 5µM MitoSOX (ThermoFisher) to measure total and mtROS, respectively. The RFUs were converted into nanomoles of ROS/mg protein, according to a titration curve performed with serial dilutions of $H_2O_2$.

Malonyl dialdehyde (MDA), an index of Lipid Peroxidation, was measured in 1×105 cells using the Lipid Peroxidation (MDA) Assay Kit (Abcam, Cambridge, UK) according to the manufacturer's instructions. The results are expressed as nmol/mg total protein. To evaluate protein oxidation, 1×105 cells were lysed and the amount of carbonylated proteins was measured using 100 µg whole cell extract with a Protein Carbonyl Content Assay Kit (Abcam). The results are expressed as nmol/mg total protein.

For glutathione measurement, 1×105 cells were washed with 480 µL PBS and proteins were precipitated by adding 120 µL of 6.5% w/v 5-sulfosalicylic acid. Each sample was placed on ice for 1 h and centrifuged for 15 min at 13,000 × g (4°C). Total glutathione was measured in 20 µL of the lysate using the following reaction mix: 20 µL stock buffer (143 mM NaH2PO4 and 63 mM EDTA, pH 7.4), 200 µL of daily reagent (10 mM 5,5’dithiobis-2-nitrobenzoic acid and 2 mM NADPH in stock buffer), and 40 µL glutathione reductase (8.5 U/ml). The content of oxidized glutathione (GSSG) was obtained after derivatization of GSH with 2-vinylpyridine (2VP); 10 µL of 2VP was added to 200 µL of lysate, and the mixture was shaken at room temperature for 1 h. Glutathione was then measured in 40 µl of sample as described. The reaction was followed kinetically for 5 min using a Synergy HT Multi-Mode Microplate Reader (Bio-Tek Instruments, Vinooski, VT, USA), and the absorbance was measured at 415 nm. Each measurement was performed in triplicate, and the results were expressed as nmol glutathione/min/mg cell protein. For each sample, reduced glutathione (GSH) was obtained by subtracting GSSG from total glutathione.

The activity of glucose 6-phosphate dehydrogenase (G6PD) was measured in cells washed with fresh medium, detached with trypsin/EDTA, resuspended at 0.1 x 106 cells/ml in 0.1 M Tris/0.5 mM EDTA pH 8.0, and sonicated on ice with two 10 s bursts. The cell lysate was supplemented with 10 mM MgCl2 and 0.25 mM NADP. The reaction was initiated at 37°C by adding 6-phosphogluconate (0.6 mM) with or without glucose 6-phosphate (0.6 mM), and the absorbance was measured spectrophotometrically at 340 nm using a Synergy HTX 96-well microplate reader. The first measurement was performed by adding to the assay system a saturating amount of both 6-phosphogluconate and glucose 6-phosphate: the rate of NADP+ reduction was the result of both G6PD and 6PGD activities. A second assay was performed with 6-phosphogluconate as a substrate; this procedure allowed the measurement of 6PGD activity alone. G6PD activity was determined by subtracting the rate of the second assay from that of the first assay (Riganti et al. 2002). The reaction kinetics were linear throughout the 5 min observation period. Enzymatic activity was expressed as nmol NADPH/min/mg of cell protein. The activity of cytosolic superoxide dismutase 1 (SOD1) and mitochondrial superoxide dismutase 2 (SOD2) was measured using 10 μg of cytosolic and mitochondrial proteins after cytosol-mitochondrial separation, as detailed in Xu et al. (Xu et al. 2021). Samples were resuspended in 100 μL PBS and incubated with 50 μM xanthine, 5 U/ml xanthine oxidase, and 1 μg/ml oxidized cytochrome c for 5 min at 37°C. The rate of cytochrome c reduction, which is inhibited by the presence of SOD, was monitored for 5 min by reading the absorbance at 550 nm using a Packard microplate reader EL340 (Bio-Tek Instruments). Results are expressed as nmol reduced cytochrome c/min/mg cytosolic or mitochondrial proteins.

The activities of glutathione reductase (GR), glutathione peroxidase (GPX) and thioredoxin reductase (TrxR) were measured with the following commercial kits (all from Abcam):

Glutathione Reductase (GR) Assay Kit, Glutathione Peroxidase Assay Kit, Thioredoxin Reductase Assay Kit, as per manufacturer's instructions. The results were expressed as enzymatic units (U)/mg cell protein, according to the titration curve of each kit (Figure 2S).

### RESULTS

#### Characterization of Extracellular Vesicles from Bone Marrow Mesenchymal Stromal Cells (MSCs) and Adipose Mesenchymal Stromal Cells (ASCs)

Extracellular vesicles (EVs) were isolated from mesenchymal stem cells (MSCs) and adipose-derived stem cells (ASCs) and subsequently characterized to evaluate their size, concentration, morphology, integrity, and presence of typical MSC, ASC, and EV protein markers (Figure 3S). Light scattering analysis revealed that MSC-EVs and ASC-EVs exhibited similar size distribution profiles. The mean diameters were 223.2 ± 3.1 nm and 249.3 ± 6.2 nm, and the modal diameters were 179.7 ± 7.8 nm and 176.9 ± 7.3 nm, respectively (Figure 3S, A and B). Concentration measurements yielded comparable results, with 2.01 x 1011 ± 1.07 x 1010 particles/ml in MSC-EVs and 2.84 x 1011 ± 5.03 x 1010 particles/ml in ASC-EVs. The surface proteins of EVs were analyzed using cytofluorimetric analysis (Figure 3S, C). Both EV types expressed the characteristic MSC markers CD105 and CD44, the typical MSC markers CD29, CD49e, and CD146, as well as the tetraspanins CD81, CD63, and CD9. Furthermore, the mesenchymal origin was confirmed by the absence of the endothelial marker CD31 and the epithelial marker CD326 (data not shown). Transmission electron microscopy (TEM) analysis demonstrated that both EV types possessed a characteristic round morphology and maintained structural integrity (Figure 3S, D).

**Figures Legend**

**Figure 1S** Graphical representation of the experimental protocol for immunofluorescence analysis.

**Figure 2S** Graphical representation of the experimental protocol for oxidative stress assessment.

**Figure 3S.** Characterization of MSC-EVs and ASC-EVs. Representative Nanoparticle tracking analysis illustrating the size distribution of MSC-EVs (A) and ASC-EVs (B). Multiplex bead-based flow cytometry assay of various surface markers; only expressed markers are presented as mean fluorescence intensity (MFI) (C). Representative Transmission electron microscopy images of MSC-EVs and ASC-EVs (D) with a scale bar of 200 nm.

**Figure 4S** The concentrations of total reactive oxygen species (ROS), mitochondrial ROS (mtROS), lipid peroxidation, carbonylated proteins, reduced glutathione (GSH), oxidized glutathione (GSSG) and the activity of antioxidant enzymes, including glucose 6-phosphate dehydrogenase (G6PD), cytosolic superoxide dismutase 1 (SOD1), mitochondrial superoxide dismutase 2 (SOD2), glutathione reductase (GR), glutathione peroxidase (GPX), and thioredoxin reductase (TrxR) were measured in BEAS-2B cells following exposure to mesenchymal stem cell-derived extracellular vesicles (MSC-EVs) and adipose-derived stem cell extracellular vesicles (ASC-EVs), either concurrently or six hours post-exposure to plasma from patients with ARDS under VV ECMO or healthy volunteers. The data are expressed as median and IQR. Individual points represent single measurements. Statistical significance was denoted as follows: *$P<0.05$, **$P<0.01$, ***$P<0.001$, ****$P<0.0001$. The plasma samples tested included those from healthy individuals ($n = 5$) and ARDS patients under VV ECMO ($n = 10$).

**Figure 5S** The concentrations of total reactive oxygen species (ROS), mitochondrial ROS (mtROS), lipid peroxidation, carbonylated proteins, reduced glutathione (GSH), oxidized glutathione (GSSG) and the activity of antioxidant enzymes, including glucose 6-phosphate dehydrogenase (G6PD), cytosolic superoxide dismutase 1 (SOD1), mitochondrial superoxide dismutase 2 (SOD2), glutathione reductase (GR), glutathione peroxidase (GPX), and thioredoxin reductase (TrxR) were measured in SAEC cells following exposure to mesenchymal stem cell-derived extracellular vesicles (MSC-EVs) and adipose-derived stem cell extracellular vesicles (ASC-EVs), either concurrently or six hours post-exposure to plasma from patients with ARDS under VV ECMO or healthy volunteers. The data are expressed as median and IQR. Individual points represent single measurements. Statistical significance was denoted as follows: *P<0.05, **P<0.01, ***P<0.001, ****P<0.0001. The plasma samples tested included those from healthy individuals (n = 5) and ARDS patients under VV ECMO (n = 10).

**Figure 6S** The concentrations of total reactive oxygen species (ROS), mitochondrial ROS (mtROS), lipid peroxidation, carbonylated proteins, reduced glutathione (GSH), oxidized glutathione (GSSG) and the activity of antioxidant enzymes, including glucose 6-phosphate dehydrogenase (G6PD), cytosolic superoxide dismutase 1 (SOD1), mitochondrial superoxide dismutase 2 (SOD2), glutathione reductase (GR), glutathione peroxidase (GPX), and thioredoxin reductase (TrxR) were measured in BEAS-2B cells following exposure to mesenchymal stem cell-derived extracellular vesicles (MSC-EVs) and adipose-derived stem cell extracellular vesicles (ASC-EVs), either concurrently or six hours post-exposure to plasma from ARDS patients with hyper-inflammatory or hypo-inflammatory phenotype, healthy volunteers or Cytomix. The data are expressed as median and IQR. Individual points represent single measurements. Statistical significance was denoted as follows: *P<0.05, **P<0.01. The plasma samples tested included those from healthy individuals (n = 5), hyper-inflammatory ARDS patients (n = 10) and hypo-inflammatory ARDS patients (n = 10).

**Figure 1S**

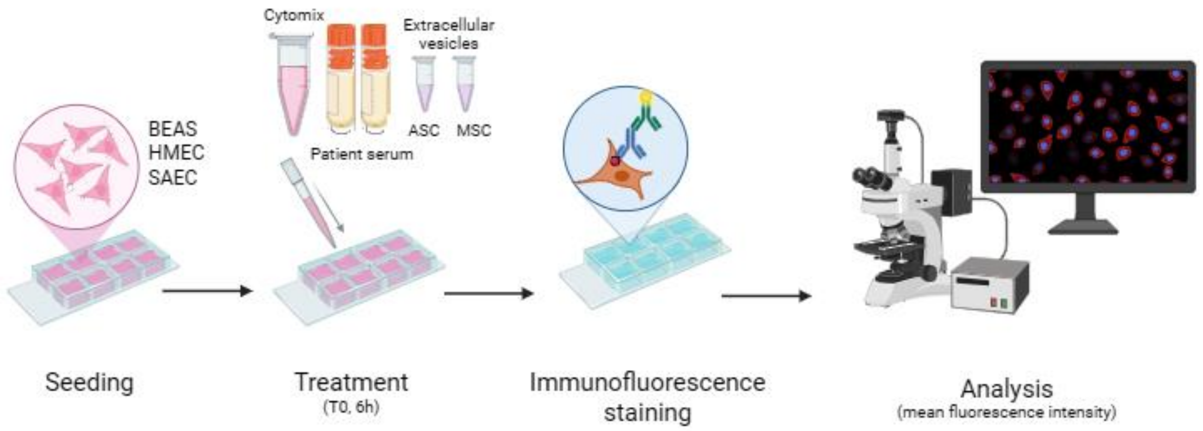


**Figure 2S**

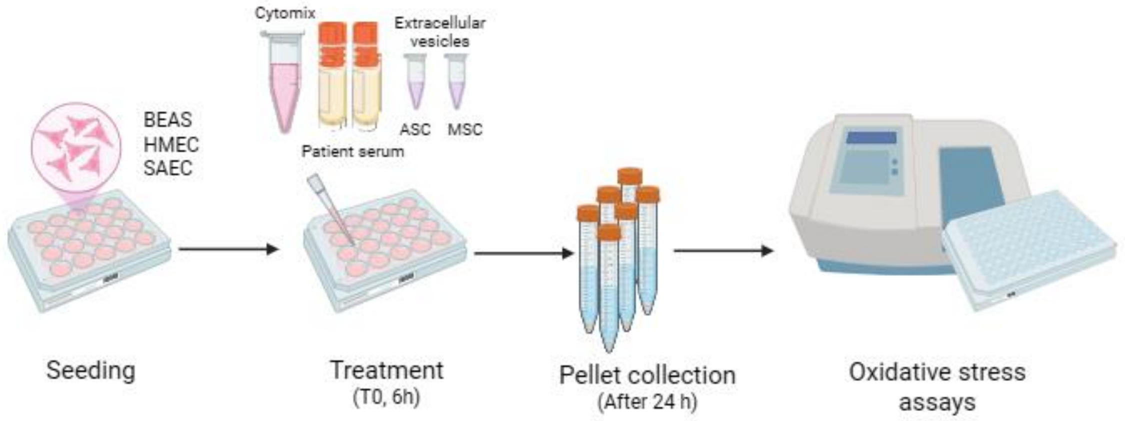

### Figure 3S

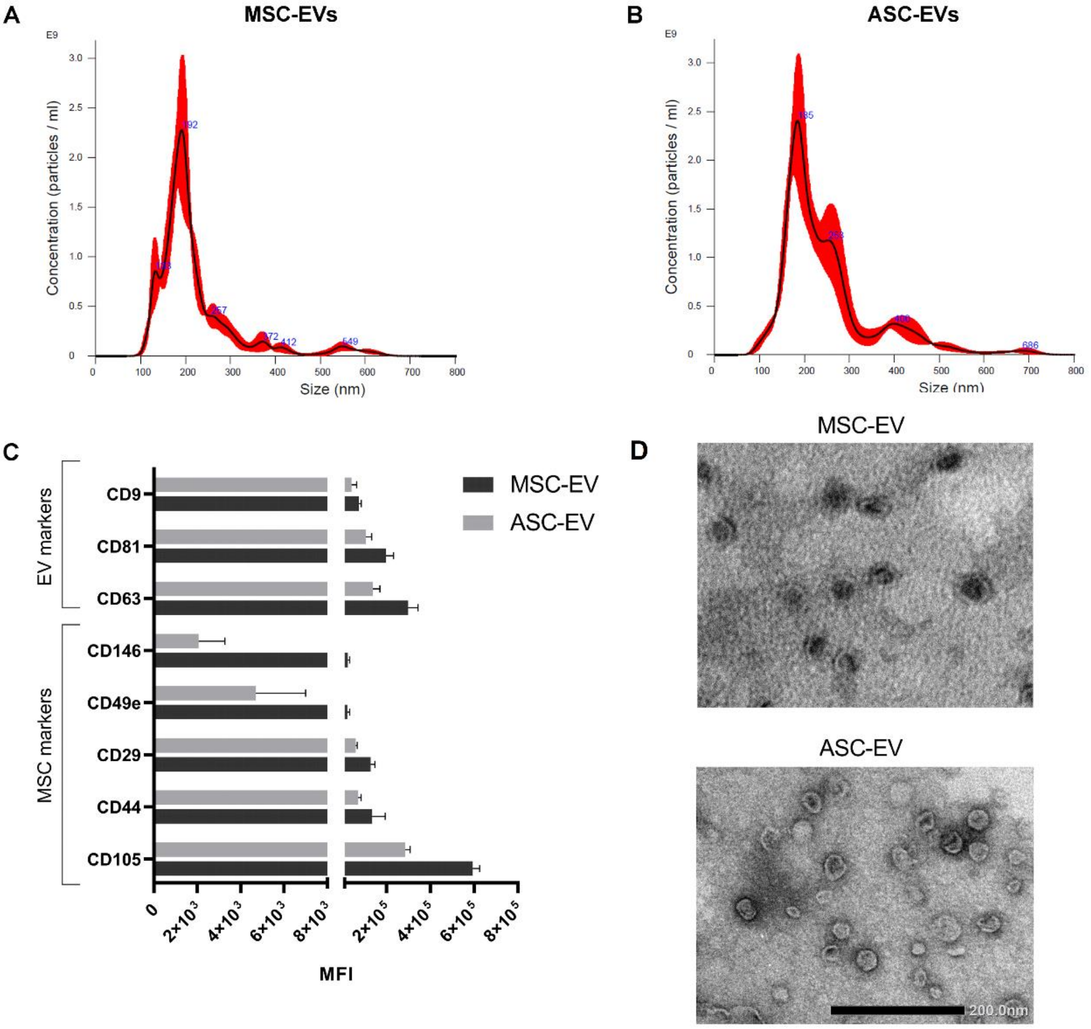

A
MSC-EVs
E9
Concentration (particles / ml)
Size (nm)
B
ASC-EVs
C
EV markers
MSC markers
CD9
CD81
CD63
CD146
CD49e
CD29
CD44
CD105
MSC-EV
ASC-EV
MFI
D
MSC-EV
ASC-EV
200.0nm

### Figure 4S

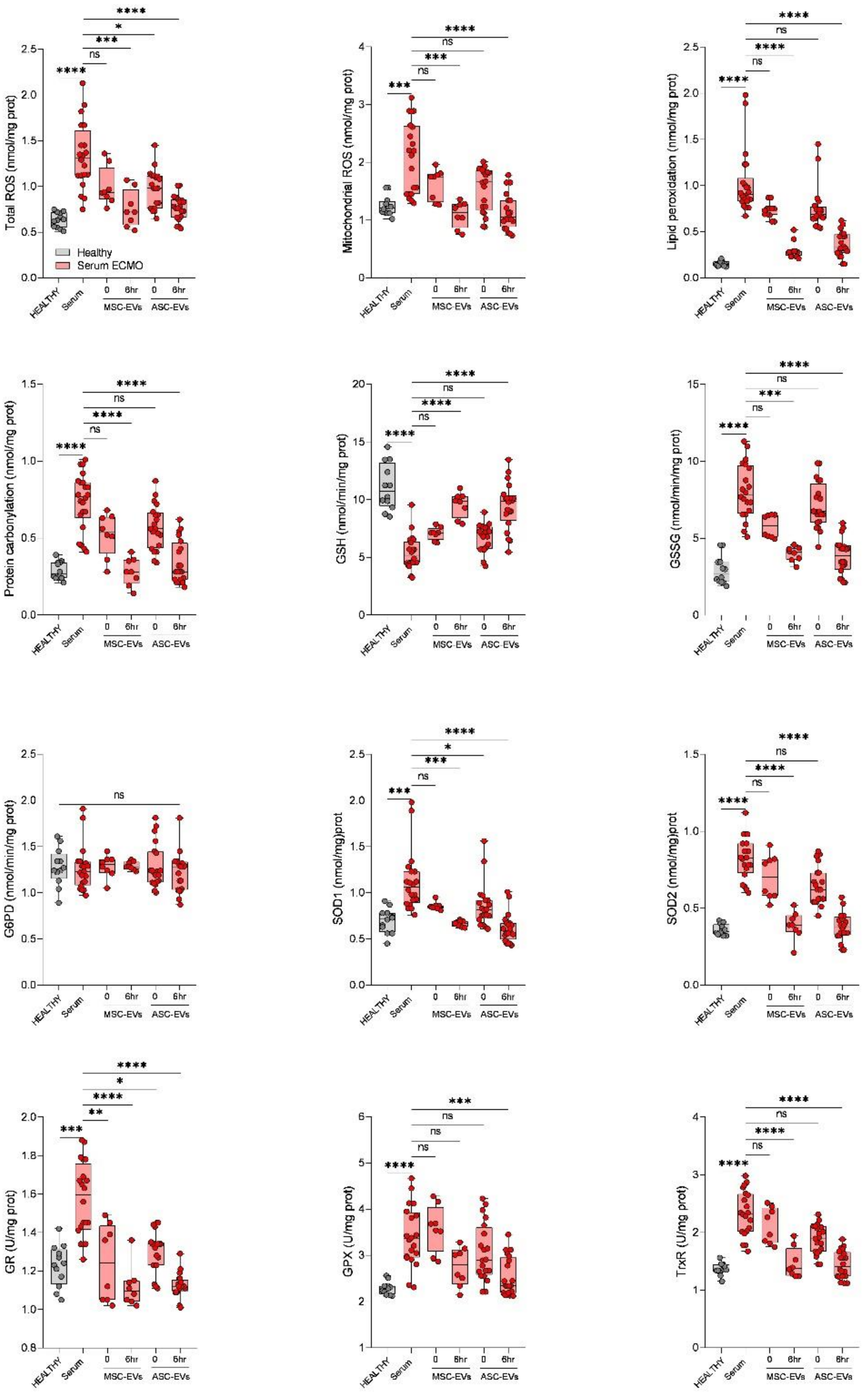

Total ROS (nmol/mg prot)
Healthy
Serum ECMO
Mitochondrial ROS (nmol/mg prot)
Lipid peroxidation (nmol/mg prot)
Protein carbonylation (nmol/mg prot)
GSH (nmol/min/mg prot)
GSSG (nmol/min/mg prot)
G6PD (nmol/min/mg prot)
SOD1 (nmol/mg)prot
SOD2 (nmol/mg)prot
GR (U/mg prot)
GPX (U/mg prot)
TrxR (U/mg prot)
HEALTHY
Serum
0
6hr
MSC-EVs
ASC-EVs

**Figure 5S**

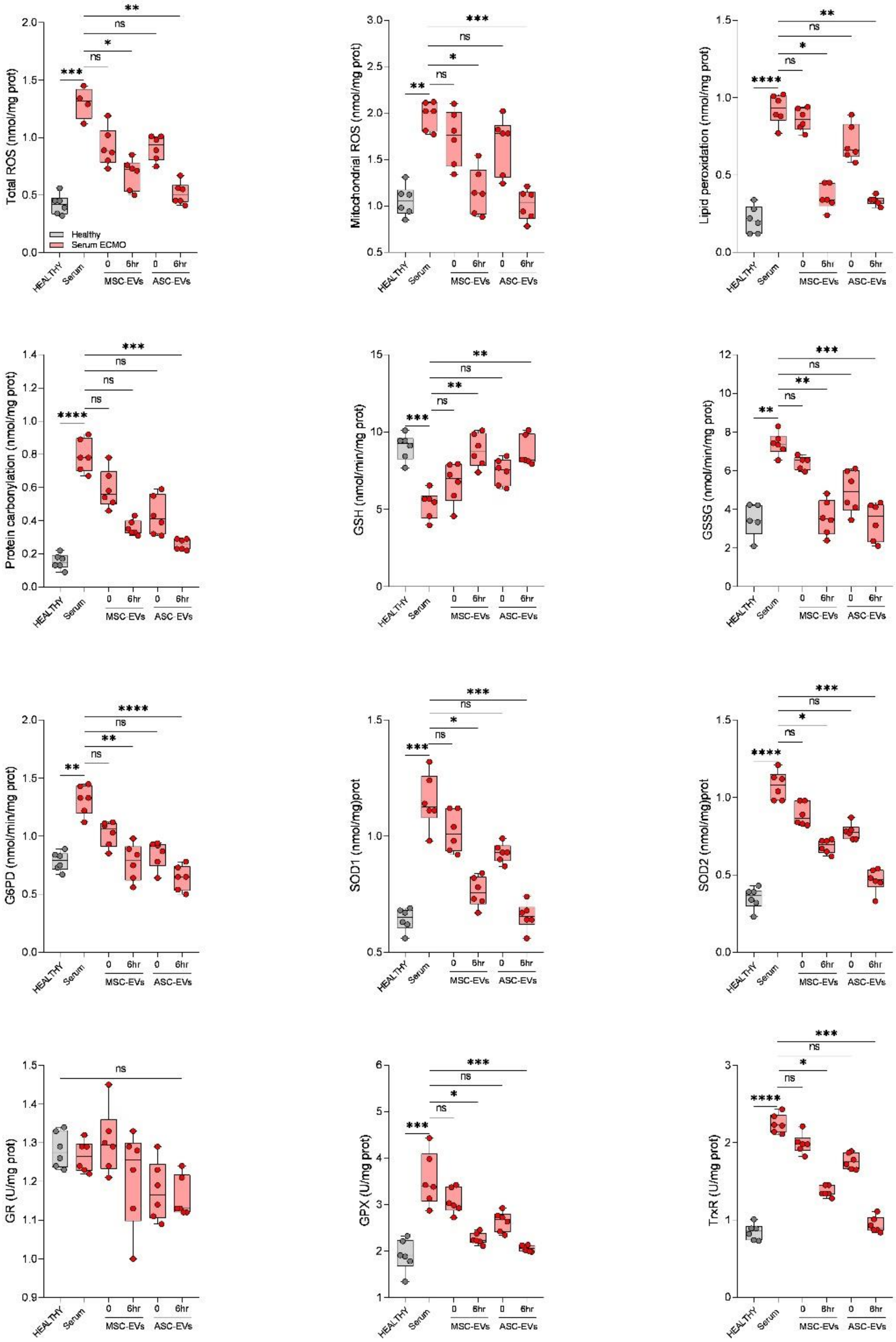
Total ROS (nmol/mg prot)
Healthy
Serum ECMO
Mitochondrial ROS (nmol/mg prot)
Lipid peroxidation (nmol/mg prot)
Protein carbonylation (nmol/mg prot)
GSH (nmol/min/mg prot)
GSSG (nmol/min/mg prot)
G6PD (nmol/min/mg prot)
SOD1 (nmol/mg)prot
SOD2 (nmol/mg)prot
GR (U/mg prot)
GPX (U/mg prot)
TrxR (U/mg prot)
HEALTHY
Serum
0
6hr
MSC-EVs
ASC-EVs

### Figure 6S

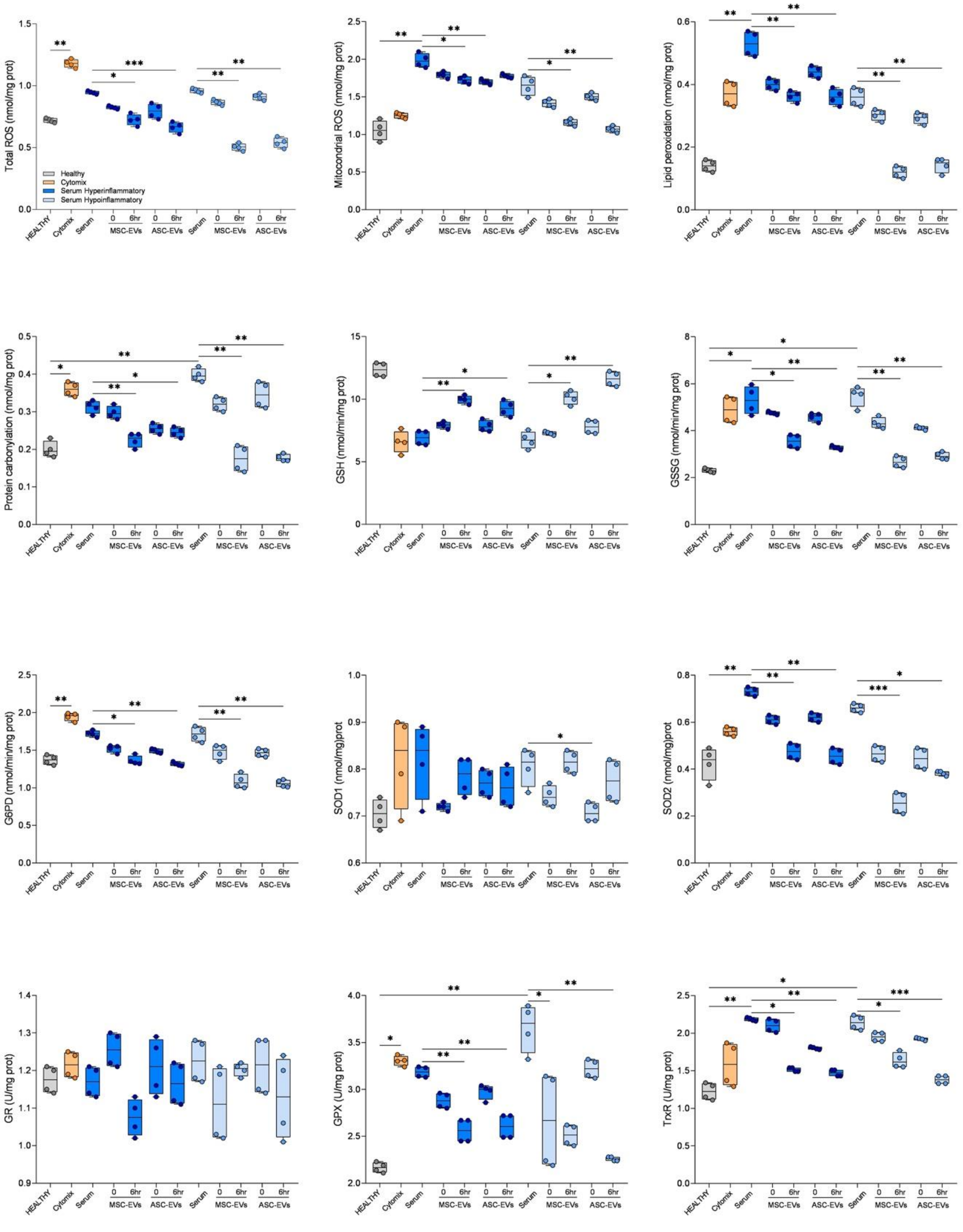
Healthy
Cytomix
Serum Hyperinflammatory
Serum Hypoinflammatory
Total ROS (nmol/mg prot)
Mitocondrial ROS (nmol/mg prot)
Lipid peroxidation (nmol/mg prot)
Protein carbonylation (nmol/mg prot)
GSH (nmol/min/mg prot)
GSSG (nmol/min/mg prot)
G6PD (nmol/min/mg prot)
SOD1 (nmol/mg)prot
SOD2 (nmol/mg)prot
GR (U/mg prot)
GPX (U/mg prot)
TrxR (U/mg prot)
HEALTHY
Cytomix
Serum
0
6hr
MSC-EVs
ASC-EVs
